# Behavioral Responses to Risk Promote Vaccinating High-contact Individuals First

**DOI:** 10.1101/2021.02.05.21251215

**Authors:** Hazhir Rahmandad

## Abstract

If COVID-19’s reproduction number was constant, vaccinating elderly first minimized deaths. However, incorporating risk-driven behavior/policy changes enhances fit to data and prioritizes vaccinating high-contact individuals. Deaths grow exponentially until people are compelled to reduce contacts, stabilizing at levels obliging higher-contact groups to sufficiently cut interactions. Vaccinating those groups out of transmission saves lives and speeds everybody’s return to normal life.

---

An analysis of alternative COVID-19 vaccination priorities by Bubar et. al. (*1*) (BL for shorthand) supports vaccinating the elderly first to minimize total deaths and years of life lost (YLL), especially if COVID-19’s basic reproduction number (*R*_*0*_) is above 1.4. Their analysis incorporates age-specific contact and fatality patterns, and vaccine effectiveness and hesitancy offering a nuanced view of vaccine prioritization under various contingencies. With elderly’s Infection Fatality Rate (IFR) orders of magnitude higher than the youth’s (*2*), the results feel intuitive as well. Yet, the analysis is missing a key factor in COVID-19 dynamics: that in response to perceived risks people and governments change their behaviors, policies, and non-pharmaceutical interventions, altering transmission rates endogenously. Such ‘*Behavioral Responses*’ to changing risk perceptions explain why despite initial *R*_*0*_ estimates much higher than one (*3*) few countries have been overrun by the disease. After an initial wave those behavioral responses brought down contacts enough for cases to oscillate around a steady flow. Most communities converge to effective reproduction numbers (*R*_*e*_) around one, otherwise deaths grow exponentially. Incorporating endogenous changes in *R*_*e*_ are important for two reasons.

First, behavioral responses are needed to explain the historical trajectory of the pandemic. Absent those, calibrating the BL model to pre-vaccination death rates in the US requires an *R*_*0*_ that peaks as low as 1.5 and still offers an unsatisfactory fit (Figure 1A). A simple extension that incorporates behavioral responses by scaling contact rates by a factor *g*(*t*) = exp^−*D*(*t*)/*α*^ matches the waves of pandemic across age groups with *R*_*0*_ peaking at 4.1 (Panel B). Here perceived risk, *D(t)*, is an exponential lag of per-million daily death rates and adjusts faster when deaths are growing (estimated lag: 8 days) than when deaths subside (lag: 42 days). Responsiveness is quantified as the risk level compelling significant contact reductions (*α*=6.9 deaths/million).

**Figure 1.**
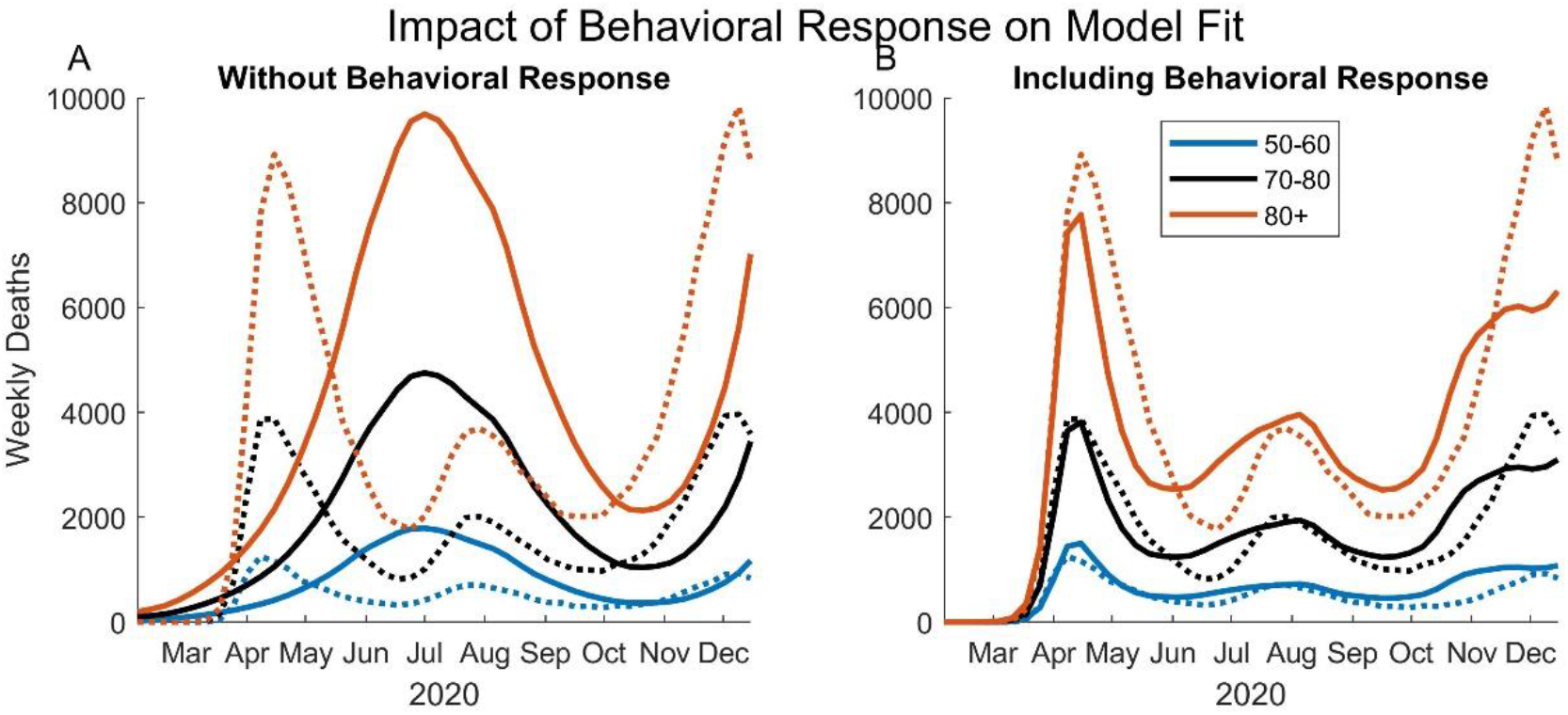
USA deaths (from CDC; dashed lines) vs. calibrated model outputs (solid lines) for three age groups. A) *R*_*0*_ and a weather effect multiplier (*4*) calibrated in the “leaky vaccine” BL model with otherwise baseline parameters. Excluding weather effect reduces fit and leads to a single-peaked contagion. B) Calibrated extended BL model with g(t).

Second, behavioral responses, also absent from most other vaccination analyses (e.g. *5, 6, 7*), change both the magnitude of deaths, and the vaccination priorities, qualitatively. Figure 2 compares alternative prioritization schemes using calibrated BL model (Panel A) and its extension ‘Including Behavioral Response’ (panel B). Consistent with BL findings *Elderly-First* policy is best when behavioral response is ignored, but incorporating this consideration flips the priority for any vaccination rollout that takes more than 6 months. Magnitude of the impact is considerable: in 2021 under calibrated BL model vaccinating the population over 250 days leads to 262 vs. 390 thousand deaths for *Elderly-First* and *High-contact-First* policies respectively (blue dot in Panel A is the ratio, 0.67). The corresponding projections accounting for behavioral responses change to 125 and 110 thousands of deaths from vaccination start to the successful suppression of epidemic (ratio of 1.14; Panel B). The large difference in deaths is due to the timing of projected epidemic waves. Absent behavioral responses the increased *R*_*e*_ due to winter weather leads to a large “second” wave in the US (Figure 1A) raising deaths significantly before vaccination can curb the pandemic. With behavioral response the “third” wave is endogenously brought under control, with under half projected deaths during vaccination period.

**Figure 2.**
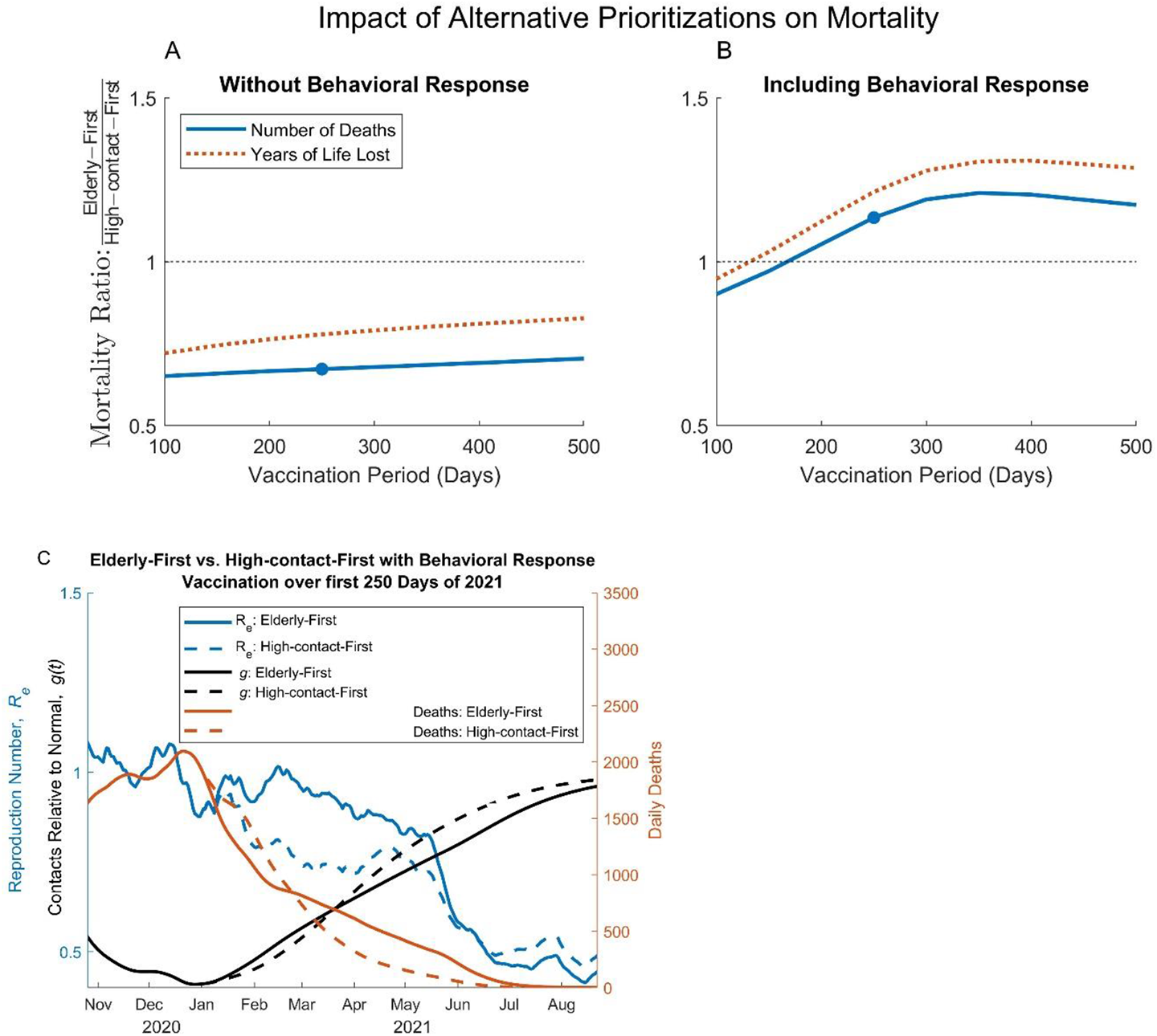
Comparing Elderly-first and High-contact-First policies. Each prioritize the vaccination among 10-year age groups based on their age, or total contact rates, respectively. Ratio of cumulative deaths (and YLL) under Elderly-First vs. High-contact-First policies from beginning of vaccination (January 2021) until full control in calibrated models. A) BL model and B) extended BL with behavioral response. C) Elderly-First (solid) vs. High-contact-First (dashed) under behavioral response. Projections of contacts relative to normal (g(t); black, left axis), Effective reproduction number (blue, left axis), and daily death rates (red, right axis) are shown. Parameters as in baseline BL: Vaccine effectiveness: 90%; vaccine hesitancy: 30%.

Exact long-term projections may not be reliable, but the rank ordering of vaccination policies is robust. That ordering switches with inclusion of behavioral response and can be understood in light of two competing mechanisms. First, vaccinating groups based on their IFR reduces deaths among infected, promoting elderly-first policy as BL recommends. The second mechanism operates through a subtler pathway: when people change their behaviors in response to risk, ongoing deaths are determined largely by how responsive to risk different groups in a society are. Deaths stabilize at levels that compel enough reductions in infectious contacts to curtail the exponential growth of the epidemic, i.e. keep *R*_*e*∼_1. Higher-contact, or less-responsive, groups would bring down their interactions sufficiently when death rates are higher (than rates tolerable to e.g. lower-contact elderly) (*8*). Thus, ongoing deaths could be brought down further by first vaccinating those with higher contact rates (e.g. the High-contact-Frist policy), or those less responsive to risks (a dimension not explicitly modeled here). Taking these groups out of transmission dynamics brings down deaths by shifting the underlying (pseudo-)equilibrium death rates implied by behavioral responses. Initially the first mechanism brings down deaths faster under Elderly-First policy (Figure 2C). However, due to the second mechanism remaining high-contact individuals keep the reproduction number higher (than High-contact-First) under Elderly-First prioritization, increasing infections and the later deaths enough to change the overall conclusion. Results are even more pronounced for YLL. Moreover, the life-saving benefits of High-contact-First policy are complemented by the faster return to normal interaction patterns (black lines).

Behavioral responses, missing from existing analysis of vaccination priority, are not only needed for understanding the observed and future trajectories of the pandemic, but also shift the optimal vaccination policy significantly in favor of prioritizing high contact groups first. Moreover, the underlying mechanism is very relevant if one could incorporate into priorities differences in risk-responsiveness across population groups. Prioritization of higher-contact, as well as those least able to change their behaviors in response to COVID-19 risks (e.g. health-care workers, first-responders, incarcerated, low-income service workers, and minority communities that have suffered the greatest COVID-19 deaths to-date) may save many thousands of lives while enabling a faster return to normal life for everybody.

## Data Availability

All models, data, and analysis code are available at: https://www.dropbox.com/s/48m5qn9bx697i8r/VaccineExample.zip?dl=0

## Acknowledgments

Navid Ghaffarzadegan, Tse Yang Lim, John Sterman, and Kim Thompson provided helpful comments.

